# Gene specific effects on brain volume and cognition of *TMEM106B* in frontotemporal lobar degeneration

**DOI:** 10.1101/2024.04.05.24305253

**Authors:** Marijne Vandebergh, Eliana Marisa Ramos, Nick Corriveau-Lecavalier, Vijay K Ramanan, John Kornak, Carly Mester, Tyler Kolander, Danielle Brushaber, Adam M Staffaroni, Daniel Geschwind, Amy Wolf, Kejal Kantarci, Tania F Gendron, Leonard Petrucelli, Marleen Van den Broeck, Sarah Wynants, Matthew C Baker, Sergi Borrego – Écija, Brian Appleby, Sami Barmada, Andrea Bozoki, David Clark, R Ryan Darby, Bradford C Dickerson, Kimiko Domoto-Reilly, Julie A. Fields, Douglas R. Galasko, Nupur Ghoshal, Neill Graff-Radford, Ian M Grant, Lawrence S Honig, Ging-Yuek Robin Hsiung, Edward D Huey, David Irwin, David S Knopman, Justin Y Kwan, Gabriel C Léger, Irene Litvan, Joseph C Masdeu, Mario F Mendez, Chiadi Onyike, Belen Pascual, Peter Pressman, Aaron Ritter, Erik D Roberson, Allison Snyder, Anna Campbell Sullivan, M Carmela Tartaglia, Dylan Wint, Hilary W Heuer, Leah K Forsberg, Adam L Boxer, Howard J Rosen, Bradley F Boeve, Rosa Rademakers, the ALLFTD Consortium

**Affiliations:** VIB Center for Molecular Neurology, VIB, Antwerp, Belgium; Department of Biomedical Sciences, University of Antwerp, Antwerp, Belgium; Department of Neurology, David Geffen School of Medicine, University of California, Los Angeles, Los Angeles, CA, USA; Department of Neurology, Mayo Clinic, Rochester, MN, USA; Department of Psychiatry and Psychology, Mayo Clinic, Rochester, MN, USA; Department of Epidemiology and Biostatistics, University of California, San Francisco, San Francisco, CA, USA; Department of Quantitative Health Sciences, Mayo Clinic, Rochester, MN, USA; Department of Neurology, Memory and Aging Center, University of California, San Francisco Weill Institute for Neurosciences, San Francisco, CA, USA; Institute for Precision Health, Departments of Neurology, Psychiatry and Human Genetics at David Geffen School of Medicine, University of California, Los Angeles, Los Angeles, CA, USA; Department of Neuroscience, Mayo Clinic, Jacksonville, FL, USA; Alzheimer’s Disease and Other Cognitive Disorders Unit, Neurology Service, Hospital Clínic de Barcelona, Institut d’Investigacions Biomèdiques August Pi i Sunyer (IDIBAPS), Fundació Clínic per a la Recerca Biomèdica, Universitat de Barcelona, Barcelona, Spain; Department of Neurology, Case Western Reserve University, Cleveland, OH, USA; Department of Neurology, University of Michigan, Ann Arbor, MI, USA; Department of Neurology, University of North Carolina, Chapel Hill, NC, USA; Department of Neurology, Indiana University, Indianapolis, IN, USA; Department of Neurology, Vanderbilt University, Nashville, TN, USA; Department of Neurology, University of Washington, Seattle, WA, USA; Department of Neurosciences, University of California, San Diego, La Jolla, CA, USA; Departments of Neurology and Psychiatry, Washington University School of Medicine, Washington University, St. Louis, MO, USA; Department of Psychiatry and Behavioral Sciences, Northwestern Feinberg School of Medicine, Chicago, IL, USA; Taub Institute for Research on Alzheimer’s Disease and the Aging Brain, College of Physicians and Surgeons, Columbia University, New York, NY, USA; Department of Neurology, Columbia University, New York, NY, USA; Division of Neurology, University of British Columbia, Vancouver, British Columbia, Canada; Department of Psychiatry and Human Behavior, Alpert Medical School of Brown University, Providence, Rhode Island, USA; Department of Neurology and Penn Frontotemporal Degeneration Center, Perelman School of Medicine, University of Pennsylvania, Philadelphia, PA, USA; National Institute of Neurological Disorders and Stroke, National Institutes of Health, Bethesda, MD, USA; Department of Neurology, Houston Methodist, Houston, TX, USA; Department of Psychiatry and Behavioral Sciences, Johns Hopkins University, Baltimore, MD, USA; Department of Neurology, University of Colorado, Aurora, CO, USA; Cleveland Clinic Lou Ruvo Center for Brain Health, Las Vegas, NV, 89106, USA; Department of Neurology, University of Alabama at Birmingham, Birmingham, AL, USA; Glenn Biggs Institute for Alzheimer’s & Neurodegenerative Diseases, UT Health San Antonio; Tanz Centre for Research in Neurodegenerative Diseases, Division of Neurology, University of Toronto, Toronto, Ontario, Canada

**Author notes:** Corresponding author: Rosa Rademakers, Ph.D. (ORCID: 0000-0002-4049-0863) VIB Center for Molecular Neurology Universiteitsplein 1, 2610 Wilrijk, Belgium.

## Abstract

**Background and Objectives:** *TMEM106B* has been proposed as a modifier of disease risk in FTLD-TDP, particularly in *GRN* mutation carriers. Furthermore, *TMEM106B* has been investigated as a disease modifier in the context of healthy aging and across multiple neurodegenerative diseases. The objective of this study is to evaluate and compare the effect of *TMEM106B* on gray matter volume and cognition in each of the common genetic FTD groups and in sporadic FTD patients.

**Methods:** Participants were enrolled through the ARTFL/LEFFTDS Longitudinal Frontotemporal Lobar Degeneration (ALLFTD) study, which includes symptomatic and presymptomatic individuals with a pathogenic mutation in *C9orf72, GRN, MAPT, VCP, TBK1, TARDBP,* symptomatic non-mutation carriers, and non-carrier family controls. All participants were genotyped for the *TMEM106B* rs1990622 SNP. Cross-sectionally, linear mixed-effects models were fitted to assess an association between *TMEM106B* and genetic group interaction with each outcome measure (gray matter volume and UDS3-EF for cognition), adjusting for education, age, sex and CDR®+NACC-FTLD sum of boxes. Subsequently, associations between *TMEM106B* and each outcome measure were investigated within the genetic group. For longitudinal modeling, linear mixed-effects models with time by *TMEM106B* predictor interactions were fitted.

**Results:** The minor allele of *TMEM106B* rs1990622, linked to a decreased risk of FTD, associated with greater gray matter volume in *GRN* mutation carriers under the recessive dosage model. This was most pronounced in the thalamus in the left hemisphere, with a retained association when considering presymptomatic *GRN* mutation carriers only. The minor allele of *TMEM106B* rs1990622 also associated with greater cognitive scores among all *C9orf72* mutation carriers and in presymptomatic *C9orf72* mutation carriers, under the recessive dosage model.

**Discussion:** We identified associations of *TMEM106B* with gray matter volume and cognition in the presence of *GRN* and *C9orf72* mutations. This further supports *TMEM106B* as modifier of TDP-43 pathology. The association of *TMEM106B* with outcomes of interest in presymptomatic *GRN* and *C9orf72* mutation carriers could additionally reflect TMEM106B’s impact on divergent pathophysiological changes before the appearance of clinical symptoms.

## INTRODUCTION

Frontotemporal lobar degeneration (FTLD) is one of the leading causes of dementia in individuals younger than 65 years and represents 10-20% of all dementias. The term frontotemporal dementia (FTD) is used as an umbrella term for the spectrum of clinical manifestations that may result from FTLD, such as progressive changes in behavior or language difficulties. Some patients may also develop amyotrophic lateral sclerosis (ALS) or parkinsonism. One-third of patients show a strong family history, with most common genetic causes of FTD being autosomal dominant mutations in the progranulin (*GRN*) gene^1,2^, the microtubule-associated protein tau (*MAPT*) gene^3^ and the chromosome 9 open reading frame 72 (*C9orf72*) gene^4^.

Apart from autosomal dominant mutations causing FTD, additional genetic risk factors have been identified. In a genome-wide association study (GWAS) for the subgroup of FTLD patients characterized by TDP-43 pathology (FTLD-TDP), *TMEM106B* was identified as a risk factor^5^. The major allele (A) of the lead variant in the *TMEM106B* locus (rs1990622) was associated with an increased risk for developing disease. Interestingly, the association with *TMEM106B* was most pronounced in the subset of FTLD-TDP patients carrying a *GRN* mutation^5^, implying that disease risk imposed by autosomal dominant mutations is also subject to genetic modifiers. In a GWAS of symptomatic *GRN* cases versus population controls, individuals carrying the minor *TMEM106B* haplotype indeed showed a 50% lower chance of developing disease symptoms as compared to *GRN* mutation carriers without the minor *TMEM106B* haplotype^6^. Several other reports support the reduced disease penetrance associated with the minor (protective) *TMEM106B* haplotype^7^, in particular in patients with *GRN* mutations^8,9^. Strikingly, an obligate *GRN* mutation carrier was still unaffected in their 80’s, and found to be a homozygous carrier of the minor *TMEM106B* haplotype^10^. This suggests that carrying two copies of the *TMEM106B* minor allele may counteract the disease-causing effects of the *GRN* pathogenic mutation. A protective effect of the minor allele of *TMEM106B* rs1990622 SNP has also been demonstrated in *C9orf72* repeat expansion carriers, though less prominent compared with *GRN* mutation carriers^11^. While this has implications for genetic counselling, genotyping *TMEM106B* in *GRN* mutation carriers in the diagnostic setting is not routinely being performed.

*TMEM106B* has also been investigated as a disease modifier in the context of healthy aging. In elderly adults, the major risk allele of rs1990622 is associated with a smaller volume of the superior temporal gyrus, especially in the left hemisphere^12^, with more advanced TDP-43 pathology at autopsy^13^, increased biological aging in the prefrontal cortex^14^, worse cognitive function^14^ and decreased neuronal proportion^15^. Moreover, in patients with FTD carrying two copies of the risk allele (AA) compared with the (AG+GG) group, lower cortical gray matter volumes in the frontal, temporal, cingulate and insula cortices were noted^16^. *TMEM106B* has also been shown to be a modulator of gray matter volume and functional network connectivity in presymptomatic mutation carriers^17–19^, and of cognitive trajectories over time among clinical FTD patients^20^. However, associations of *TMEM106B* with structural imaging and cognition within different FTD genetic groups remains to be investigated. Beyond FTLD, *TMEM106B* has been implicated in TDP-43 pathology in Alzheimer’s disease (AD)^21^, cognition in Parkinson’s disease (PD)^20^ and ALS, though with conflicting findings in terms of directionality of effects in ALS^22,23^.

In this study, we aimed to investigate the modifying effects of *TMEM106B* in the largest collection of systematically ascertained FTD patients and families from the ARTFL/LEFFTDS Longitudinal Frontotemporal Lobar Degeneration (ALLFTD) study, on gray matter volume and cognitive measures. Understanding the modifying effects of *TMEM106B* across genetic FTD subtypes is crucial in light of genetic counselling and the development of gene-based therapies.

## MATERIAL AND METHODS

### Study participants and genetic analysis

Participants were enrolled through Advancing Research and Treatment for Frontotemporal Lobar Degeneration (ARTFL, NCT02365922) and Longitudinal Evaluation of Familial Frontotemporal Dementia Subjects (LEFFTDS, NCT02372773)^24^ which combined into the ARTFL/LEFFTDS Longitudinal Frontotemporal Lobar Degeneration (ALLFTD, NCT04363684) study. These studies enrolled participants through a consortium of 27 centers across the US and Canada between 2015 and 2023. Here, we report data from the most recent study visits for each participant as of October 26, 2023. This study involves human participants and was approved by Johns Hopkins Medicine Institutional Review Board (IRB) serving as the single IRB for the ALLFTD Consortium (CR00042454/IRB00227492). All participants provided written informed consent or assent with proxy consent.

ALLFTD participants had genetic testing at the University of California, Los Angeles using published methods^25^. Briefly, DNA samples were screened for genes previously implicated in neurodegenerative diseases, including *GRN*, *MAPT, TBK1, VCP, TARDBP,* using targeted sequencing or whole-exome sequencing. The presence of hexanucleotide repeat expansions in *C9orf72* was detected using both fluorescent and repeat-primed PCR. *TMEM106B* rs1990622 genotyping was carried out by real-time PCR on a LightCycler® 480 System using Taqman SNP Genotyping Assays (#C 11171598_20). Assays were run in duplicate.

Genome-wide SNP genotyping data from 1,975 ALLFTD participants was used to perform lineage analysis using PLINK. Briefly, QC was performed to remove individuals with low call rate and filter autosomal SNPs for missingness, frequency, and deviation from Hardy-Weinberg equilibrium. Genetic ancestry was inferred by projecting genotyped samples into the principal components of the 1000 Genomes reference panel, using R package bigsnpr. Identity-by-descent (IBD) estimates were then calculated to determine relatedness, followed by family-network identification and pedigree reconstruction using PRIMUS^26^.

After additional filtering on the availability of clinical data (clinical phenotype, age at visit) and genetic data (mutation in *C9orf72*, *GRN*, *MAPT*, *VCP*, *TBK1*, *TARDBP* or non-carrier) a total of 1,798 individuals were retained (**Table 1**). For affected non-mutation carriers, we only retained those with an FTD spectrum disorder, defined as either behavioral variant FTD (bvFTD), FTD with amyotrophic lateral sclerosis (FTD-ALS), corticobasal syndrome (CBS), progressive supranuclear palsy (PSP), agrammatic/nonfluent primary progressive aphasia (nfvPPA) or semantic variant PPA (svPPA).

**Table 1.**
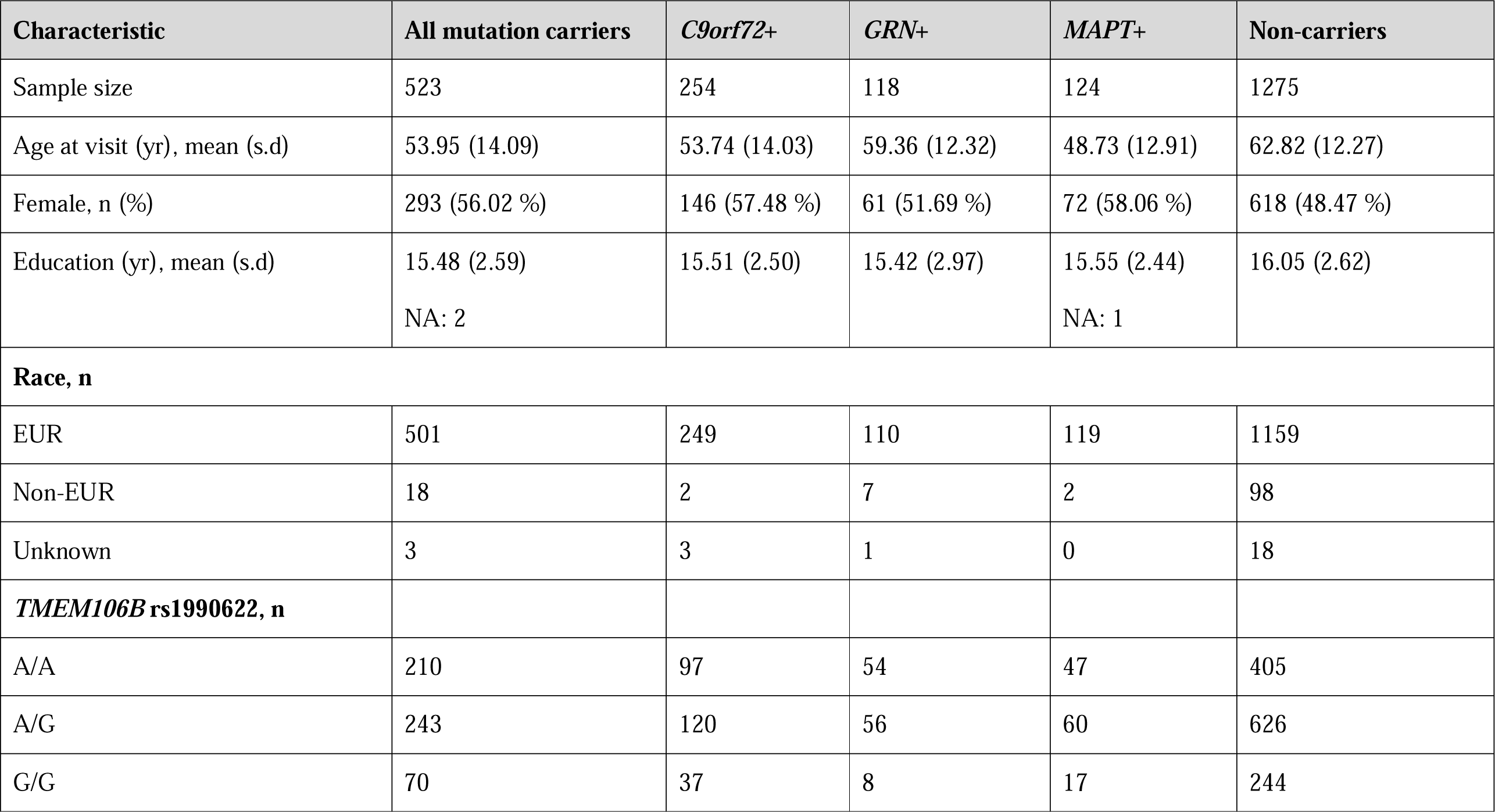
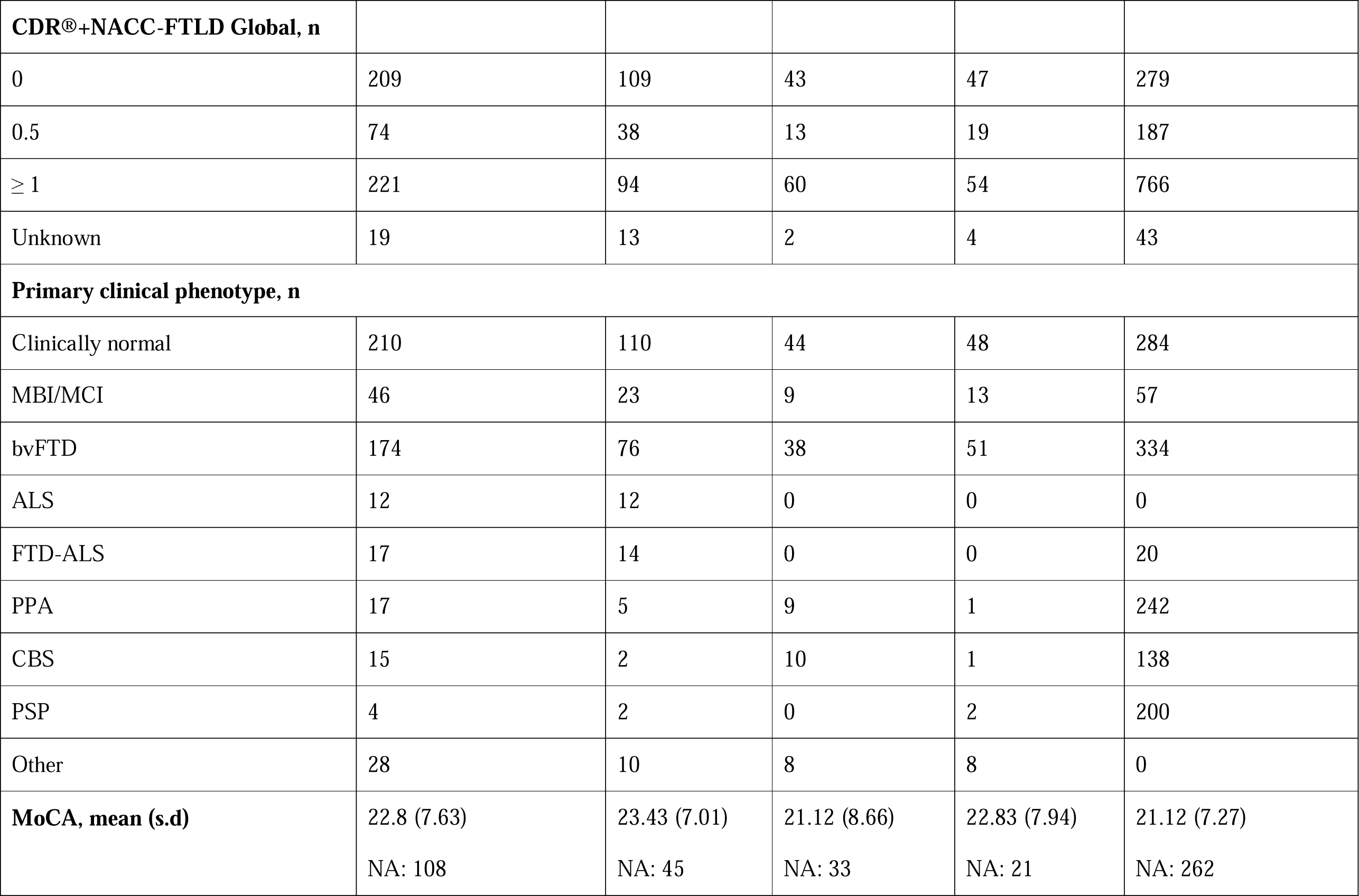
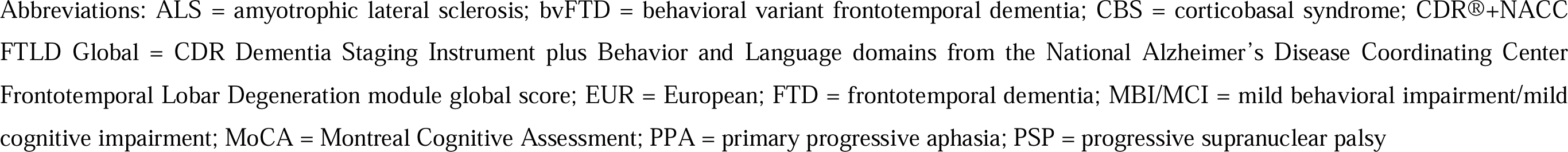
Demographic characteristics for ALLFTD participants (N = 1798)

| Characteristic | All mutation carriers | *C9orf72*+ | *GRN*+ | *MAPT*+ | Non-carriers |
| --- | --- | --- | --- | --- | --- |
| Sample size | 523 | 254 | 118 | 124 | 1275 |
| Age at visit (yr), mean (s.d) | 53.95 (14.09) | 53.74 (14.03) | 59.36 (12.32) | 48.73 (12.91) | 62.82 (12.27) |
| Female, n (%) | 293 (56.02 %) | 146 (57.48 %) | 61 (51.69 %) | 72 (58.06 %) | 618 (48.47 %) |
| Education (yr), mean (s.d) | 15.48 (2.59)<br>NA: 2 | 15.51 (2.50) | 15.42 (2.97) | 15.55 (2.44)<br>NA: 1 | 16.05 (2.62) |
| **Race, n** |  |  |  |  |  |
| EUR | 501 | 249 | 110 | 119 | 1159 |
| Non-EUR | 18 | 2 | 7 | 2 | 98 |
| Unknown | 3 | 3 | 1 | 0 | 18 |
| ***TMEM106B* rs1990622, n** |  |  |  |  |  |
| A/A | 210 | 97 | 54 | 47 | 405 |
| A/G | 243 | 120 | 56 | 60 | 626 |
| G/G | 70 | 37 | 8 | 17 | 244 |
| **CDR®+NACC-FTLD Global, n** |  |  |  |  |  |
| 0 | 209 | 109 | 43 | 47 | 279 |
| 0.5 | 74 | 38 | 13 | 19 | 187 |
| ≥ 1 | 221 | 94 | 60 | 54 | 766 |
| Unknown | 19 | 13 | 2 | 4 | 43 |
| **Primary clinical phenotype, n** |  |  |  |  |  |
| Clinically normal | 210 | 110 | 44 | 48 | 284 |
| MBI/MCI | 46 | 23 | 9 | 13 | 57 |
| bvFTD | 174 | 76 | 38 | 51 | 334 |
| ALS | 12 | 12 | 0 | 0 | 0 |
| FTD-ALS | 17 | 14 | 0 | 0 | 20 |
| PPA | 17 | 5 | 9 | 1 | 242 |
| CBS | 15 | 2 | 10 | 1 | 138 |
| PSP | 4 | 2 | 0 | 2 | 200 |
| Other | 28 | 10 | 8 | 8 | 0 |
| **MoCA, mean (s.d)** | 22.8 (7.63) | 23.43 (7.01) | 21.12 (8.66) | 22.83 (7.94) | 21.12 (7.27) |
|  | NA: 108 | NA: 45 | NA: 33 | NA: 21 | NA: 262 |

Abbreviations: ALS = amyotrophic lateral sclerosis; bvFTD = behavioral variant frontotemporal dementia; CBS = corticobasal syndrome; CDR®+NACC FTLD Global = CDR Dementia Staging Instrument plus Behavior and Language domains from the National Alzheimer's Disease Coordinating Center Frontotemporal Lobar Degeneration module global score; EUR = European; FTD = frontotemporal dementia; MBI/MCI = mild behavioral impairment/mild cognitive impairment; MoCA = Montreal Cognitive Assessment; PPA = primary progressive aphasia; PSP = progressive supranuclear palsy

### Data collection of outcome measures

#### Neuroimaging outcome: gray matter volume

Image acquisition and processing were conducted as described previously^27^. Before any preprocessing of the images, all T1-weighted images were visually inspected for quality control. Images with excessive motion or image artifact were excluded. T1-weighted images underwent bias field correction using the N3 algorithm^28^. The segmentation was performed using SPM12 (Wellcome Trust Center for Neuroimaging, London, UK, http://www.fil.ion.ucl.ac.uk/spm) unified segmentation^29^. A customized group template was generated from the segmented gray and white matter tissues and cerebrospinal fluid by non-linear registration template generation using the Large Deformation Diffeomorphic Metric Mapping framework^30^. Subjects’ native space gray and white matter were geometrically normalized to the group template, modulated, and then smoothed in the group template. The applied smoothing used a Gaussian kernel with 8∼mm full width half maximum. Every step of the transformation was carefully inspected from the native space to the group template. Regional volume estimates were calculated from individual subjects’ smoothed, modulated gray matter in template space, by taking the mean of all voxels in several *a priori* regions of interest (ROIs)^31^. The ROIs are summarized in **eTable 1.** All measures were expressed as a percentage of total intracranial volume. For gray matter volumetric measures, data was available for 958 participants (**eTable 2**).

#### Cognitive outcome

Cognition was defined using the National Alzheimer’s Coordinating Center Uniform Data Set (v3.0) executive function composite score (UDS3-EF), as described previously^32,33^. The UDS3-EF is an item response theory-based composite derived from 7 total UDS3-EF test scores: category fluency (animals and vegetables; total correct), lexical fluency (F and L words; total correct), number span backward (total correct trials), Trail Making Test parts A and B (correct lines per minute). The cognitive domains that factor into the UDS3-EF score (e.g., mental set-shifting, verbal fluency, attention/working memory) support the potential utility as a single composite measure of the common cognitive changes observed across a range of clinical phenotypes in FTD spectrum patients. The UDS3-EF composite score was available for 1,581 participants (**eTable 3**).

### Neurofilament light chain concentrations

Plasma neurofilament concentrations were determined as described previously^34,35^. Venous blood was collected in ethylenediaminetetraacetic acid–containing tubes. Blood was centrifuged at 1500 g and 4 °C for 15 minutes. Plasma samples were aliquoted in 1000-µL polypropylene tubes and stored at −80 °C, until further use. Samples (1 thaw only) were gradually brought to room temperature before analysis. Neurofilament light chain (NfL) concentrations were quantified in duplicate using the ultrasensitive HDX analyzer by single-molecule array (Simoa) technology (Quanterix) by investigators blinded to clinical group allocation.

### Statistical analysis

All analyses were conducted in R (version 4.2.2). Linear mixed effects analyses were conducted with the function ‘lmer’ in the R package ‘lme4’ (version 1.1.31).

For all cross-sectional analyses, the last available visit with the outcome measure available was used. Linear mixed models were fitted for the assessment of the main effect of the genetic groups according to their affection status (symptomatic/asymptomatic) on outcome variables, with individuals grouped by genetic status and affection status (**eTable 4**), with education, sex, age at visit and CDR®+NACC-FTLD sum of boxes score as fixed covariates and pedigree as random effect. Due to sample size limitations (< 10), only non-mutation carriers and individuals with a mutation in *C9orf72*, *GRN* or *MAPT* were considered.

To investigate the effect of the *TMEM106B* rs1990622 genotype on gray matter volume and cognition, linear mixed models were fitted with education, age, sex, genetic status and the CDR®+NACC-FTLD sum of boxes as covariates. The statistical analyses were performed under an additive (AA vs. AG vs. GG) and recessive [(AA+AG) vs. GG] genetic model, where A and G are the major and minor allele, respectively. Secondary subgroup analyses were conducted in affected individuals only, a participant was defined as affected when the primary clinical phenotype was different from ‘clinically normal’.

In addition, the effect of *TMEM106B* genotype on gray matter volume and cognition was assessed in linear mixed effect models with interaction testing between the *TMEM106B* genotype and genetic group (non-carrier, *GRN*, *MAPT* or *C9orf72*). If *p* < 0.05 for the interaction term *TMEM106B*\*genetic group, linear mixed models were fitted for the individuals belonging to that genetic group respectively (subgroup analyses), with education, age at visit, sex and CDR®+ NACC-FTLD sum of boxes as covariate.

In longitudinal models, we used linear mixed effects models with random slopes and intercepts [(time since baseline | participant ID) + (1 | pedigree ID)] to evaluate the association between *TMEM106B* genotype dosage and longitudinal changes in gray matter volume and cognition. Each participant’s baseline was defined as the first study visit with available imaging and cognitive data. Only participants with at least two timepoints and with at least one visit with a clinical phenotype different from clinically normal were included. To determine whether *TMEM106B* genotype dosages were associated with rates of change in clinical outcomes, we examined the interaction between *TMEM106B* genotype dosage and time since baseline visit, adjusting for baseline age, sex, education and baseline CDR®+NACC-FTLD sum of boxes. In addition, each genetic group was analyzed in separate models.

For the analyses with the gray matter volumes as outcome, the primary analysis was conducted with the total gray matter volume as outcome. If *p* < 0.05 for the association of *TMEM106B* genotype with total gray matter volume, secondary analyses with the individual ROIs were conducted. Sensitivity analyses were conducted excluding individuals with non-European ancestry.

### Data availability

De-identified human/patient clinical, demographic, imaging and plasma NfL data are available from ALLFTD upon request. Investigators are required to complete the Request Clinical Data form on the request portal (https://www.allftd.org/data) and to review the data sharing and publication policy. Data that could identify a participant are not provided. Any additional information required to reanalyze the data reported in this paper is available from the lead contact and ALLFTD.

## RESULTS

### Association of genetic group and affection status with gray matter volume and cognition

First, we investigated the association between the gene-affection status (combined mutation and affection status) and our outcomes of interest: total gray matter volume and cognition (defined by UDS3-EF composite score), adjusting for education, age at visit, sex, CDR®+NACC-FTLD sum of boxes. As expected, being symptomatic, regardless of genetic status, was associated with lower total gray matter volumes and lower UDS3-EF scores (**eTable 5**). In addition, being a presymptomatic *C9orf72* mutation carrier was associated with lower total gray matter volumes (beta = −1.99, 95% CI [-2.80,-1.19], *p* = 1.68 × 10^-6^) compared to clinically normal non-mutation carriers (**eTable 5**).

### Association of *TMEM106B* rs1990622 with gray matter volume

Next, we investigated the association between *TMEM106B* rs1990622 and total gray matter volume in the complete cohort, including sporadic and genetic FTD patients, presymptomatic carriers and non-mutation carrier controls. In linear mixed models with genetic status, years of education, sex, age at visit and CDR®+ NACC-FTLD sum of boxes score as fixed covariates and pedigree as random effect, *TMEM106B* rs1990622 did not statistically associate with total gray matter volume with our sample sizes, neither in the additive dosage model nor in the recessive model (**eTable 6)**. In subgroup analyses in all affected individuals, including sporadic and genetic FTD, *TMEM106B* rs1990622 did also not statistically associate with total gray matter volume (**eTable 7**) (p > 0.05).

Fitting the linear mixed-interaction model between *TMEM106B* rs1990622 and genetic group (non-mutation carrier, *GRN*, *MAPT* or *C9orf72*), with fixed covariates: years of education, sex, age at visit and CDR®+NACC-FTLD sum of boxes and with pedigree as random effect, a protective effect of the minor allele of *TMEM106B* rs1990622 on total gray matter volume was observed with additive and recessive *TMEM106B* dosages in interaction analyses with *GRN* (**Table 2**). In both the additive and recessive model, statistically significant protective effects on the gray matter volumes of the right caudal anterior cingulate, right cerebellum, left rostral caudal anterior cingulate and left frontal cortex were observed (**Table 3**). In the recessive model, the most significantly associated region was the left thalamus (*p* < 9.05 × 10^-5^, **Table 3**).

**Table 2.**
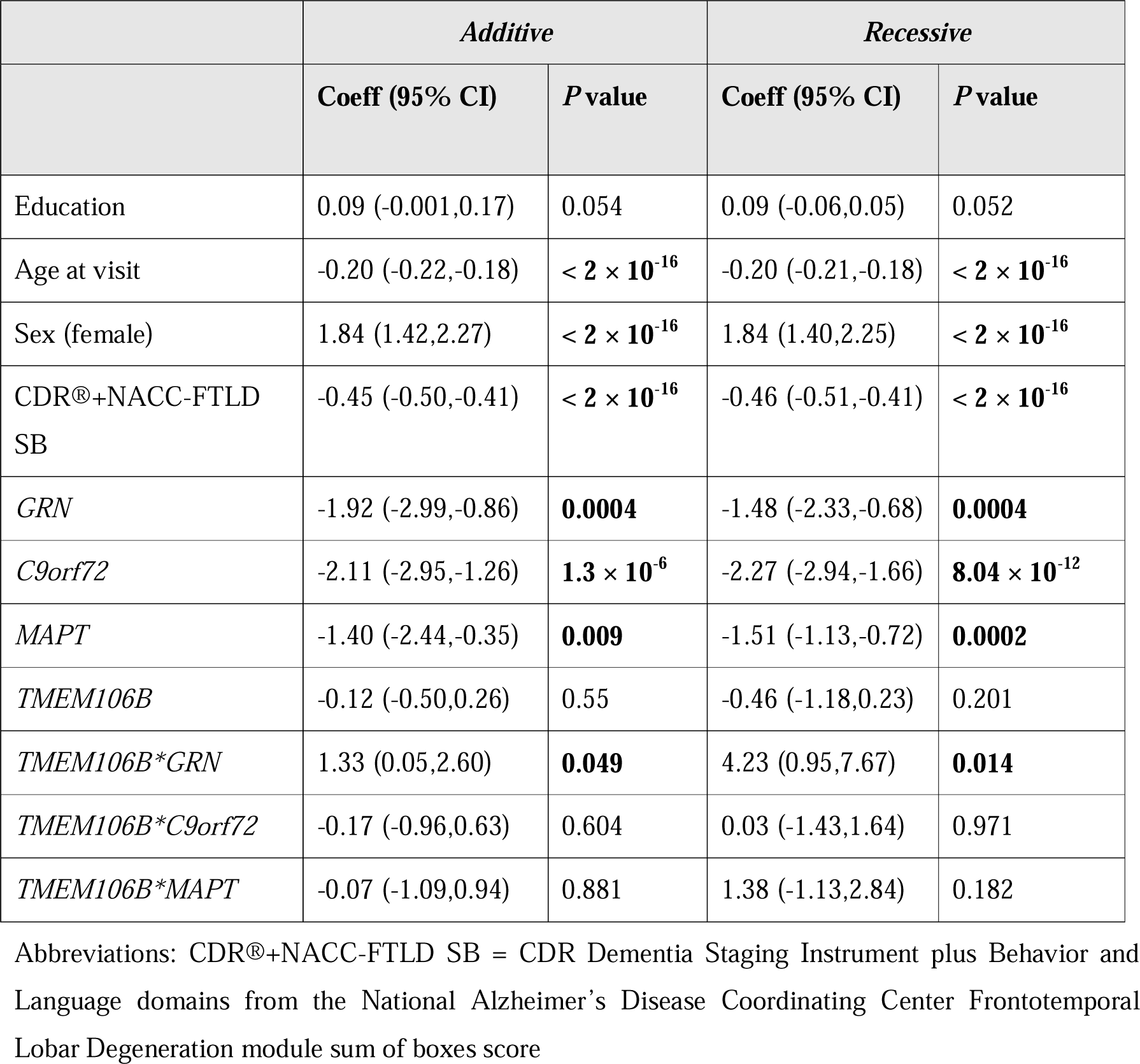
Linear mixed model statistics for *TMEM106* rs1990622 by genetic group interaction on total gray matter volume.

|  | *Additive* |  | *Recessive* |  |
| --- | --- | --- | --- | --- |
|  | **Coeff (95% CI)** | **P value** | **Coeff (95% CI)** | **P value** |
| Education | 0.09 (-0.001,0.17) | 0.054 | 0.09 (-0.06,0.05) | 0.052 |
| Age at visit | -0.20 (-0.22,-0.18) | $< 2 \times 10^{-16}$ | -0.20 (-0.21,-0.18) | $< 2 \times 10^{-16}$ |
| Sex (female) | 1.84 (1.42,2.27) | $< 2 \times 10^{-16}$ | 1.84 (1.40,2.25) | $< 2 \times 10^{-16}$ |
| CDR®+NACC-FTLD SB | -0.45 (-0.50,-0.41) | $< 2 \times 10^{-16}$ | -0.46 (-0.51,-0.41) | $< 2 \times 10^{-16}$ |
| *GRN* | -1.92 (-2.99,-0.86) | **0.0004** | -1.48 (-2.33,-0.68) | **0.0004** |
| *C9orf72* | -2.11 (-2.95,-1.26) | $1.3 \times 10^{-6}$ | -2.27 (-2.94,-1.66) | $8.04 \times 10^{-12}$ |
| *MAPT* | -1.40 (-2.44,-0.35) | **0.009** | -1.51 (-1.13,-0.72) | **0.0002** |
| *TMEM106B* | -0.12 (-0.50,0.26) | 0.55 | -0.46 (-1.18,0.23) | 0.201 |
| *TMEM106B\*GRN* | 1.33 (0.05,2.60) | **0.049** | 4.23 (0.95,7.67) | **0.014** |
| *TMEM106B\*C9orf72* | -0.17 (-0.96,0.63) | 0.604 | 0.03 (-1.43,1.64) | 0.971 |
| *TMEM106B\*MAPT* | -0.07 (-1.09,0.94) | 0.881 | 1.38 (-1.13,2.84) | 0.182 |

Abbreviations: CDR®+NACC-FTLD SB = CDR Dementia Staging Instrument plus Behavior and Language domains from the National Alzheimer's Disease Coordinating Center Frontotemporal Lobar Degeneration module sum of boxes score

**Table 3.**
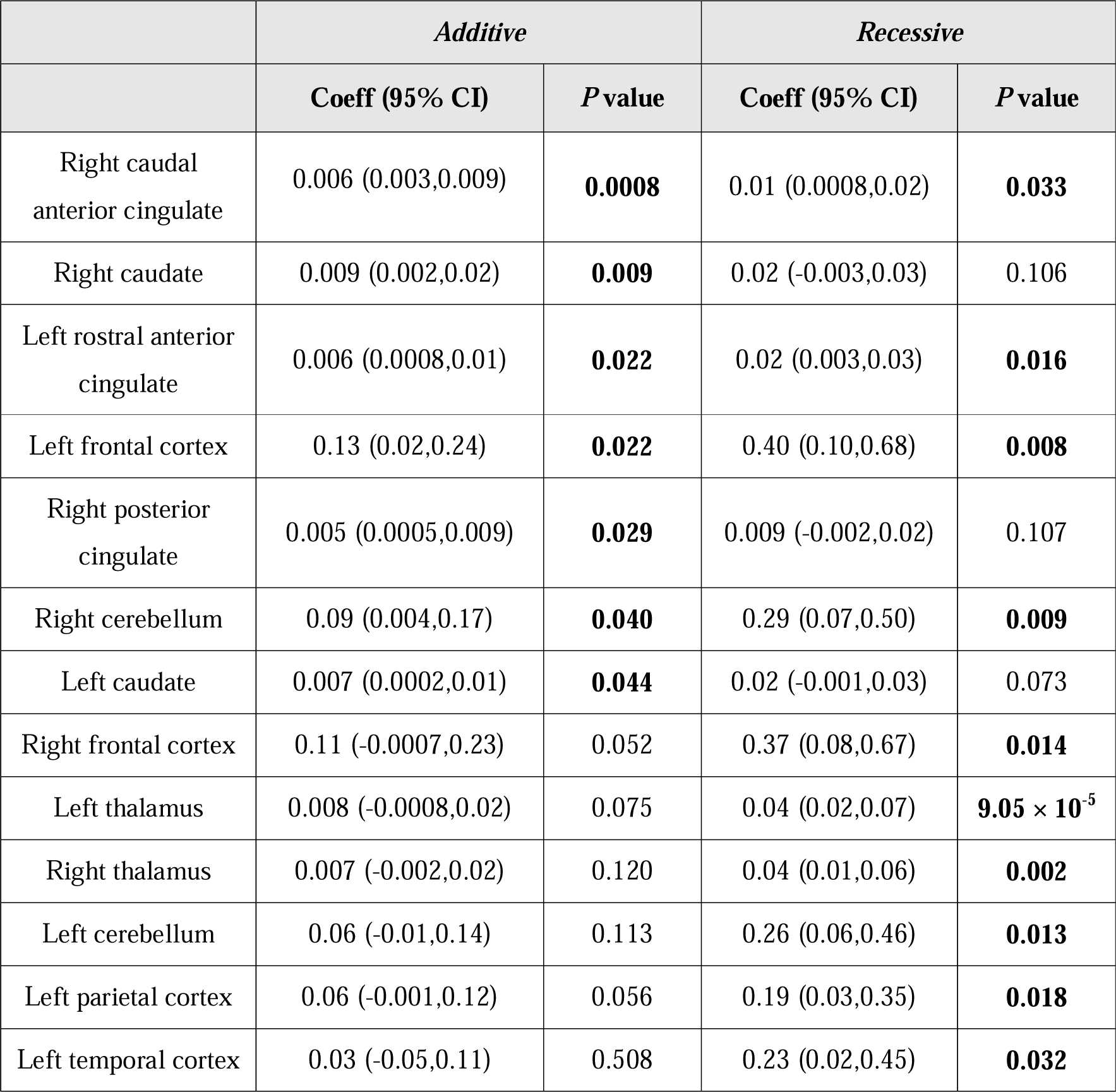
Linear mixed model statistics for *TMEM106B* rs1990622\**GRN* interaction on individual gray matter regions. Results are depicted for regions with *p* < 0.05 for either the additive or recessive *TMEM106B* genotype dosage\**GRN* interaction

|  | *Additive* |  | *Recessive* |  |
| --- | --- | --- | --- | --- |
|  | **Coeff (95% CI)** | **P value** | **Coeff (95% CI)** | **P value** |
| Right caudal anterior cingulate | 0.006 (0.003,0.009) | **0.0008** | 0.01 (0.0008,0.02) | **0.033** |
| Right caudate | 0.009 (0.002,0.02) | **0.009** | 0.02 (-0.003,0.03) | 0.106 |
| Left rostral anterior cingulate | 0.006 (0.0008,0.01) | **0.022** | 0.02 (0.003,0.03) | **0.016** |
| Left frontal cortex | 0.13 (0.02,0.24) | **0.022** | 0.40 (0.10,0.68) | **0.008** |
| Right posterior cingulate | 0.005 (0.0005,0.009) | **0.029** | 0.009 (-0.002,0.02) | 0.107 |
| Right cerebellum | 0.09 (0.004,0.17) | **0.040** | 0.29 (0.07,0.50) | **0.009** |
| Left caudate | 0.007 (0.0002,0.01) | **0.044** | 0.02 (-0.001,0.03) | 0.073 |
| Right frontal cortex | 0.11 (-0.0007,0.23) | 0.052 | 0.37 (0.08,0.67) | **0.014** |
| Left thalamus | 0.008 (-0.0008,0.02) | 0.075 | 0.04 (0.02,0.07) | **$9.05 \times 10^{-5}$** |
| Right thalamus | 0.007 (-0.002,0.02) | 0.120 | 0.04 (0.01,0.06) | **0.002** |
| Left cerebellum | 0.06 (-0.01,0.14) | 0.113 | 0.26 (0.06,0.46) | **0.013** |
| Left parietal cortex | 0.06 (-0.001,0.12) | 0.056 | 0.19 (0.03,0.35) | **0.018** |
| Left temporal cortex | 0.03 (-0.05,0.11) | 0.508 | 0.23 (0.02,0.45) | **0.032** |

In subgroup analyses in *GRN* mutation carriers, *TMEM106B* remained associated with the total gray matter volume in the recessive model (beta = 3.25, 95% CI [0.37,6.19], *p* = 0.034), with the left thalamic region as individual region of interest with the highest association (beta = 0.03, 95% CI [0.01-0.060], *p* = 0.006) **(eTable 8)**. Excluding the non-European *GRN* mutation carriers, *TMEM106B* remained associated with the total gray matter volume and left thalamic gray matter volume (beta = 3.44, 95% CI [0.72, 6.23], *p* = 0.018 and beta = 0.03, 95% CI [0.01, 0.06], *p* = 0.006, respectively).

*GRN* mutation carriers with the *TMEM106B* rs1990622*GG genotype are presymptomatic mutation carriers (**Figure 1**). Therefore, exploratory analyses were conducted that include only presymptomatic *GRN* mutation carriers. *TMEM106B* remained associated with the total gray matter volume (beta = 3.20, 95% CI [0.80,5.68], *p* = 0.016) and left thalamic gray matter volume (beta = 0.03, 95% CI [0.01,0.05], *p* = 0.003) in presymptomatic *GRN* mutation carriers in the recessive model after controlling for years of education, sex and age at visit (**eTable 9**). Excluding the non-European *GRN* presymptomatic individual did not materially affect the findings with observed estimates of beta = 3.16, 95% CI [0.73, 5.68], *p* = 0.018 and beta = 0.03, 95% CI [0.01, 0.05], *p* = 0.003 for the total gray matter volume and left thalamic gray matter volume respectively.

**Figure 1.**
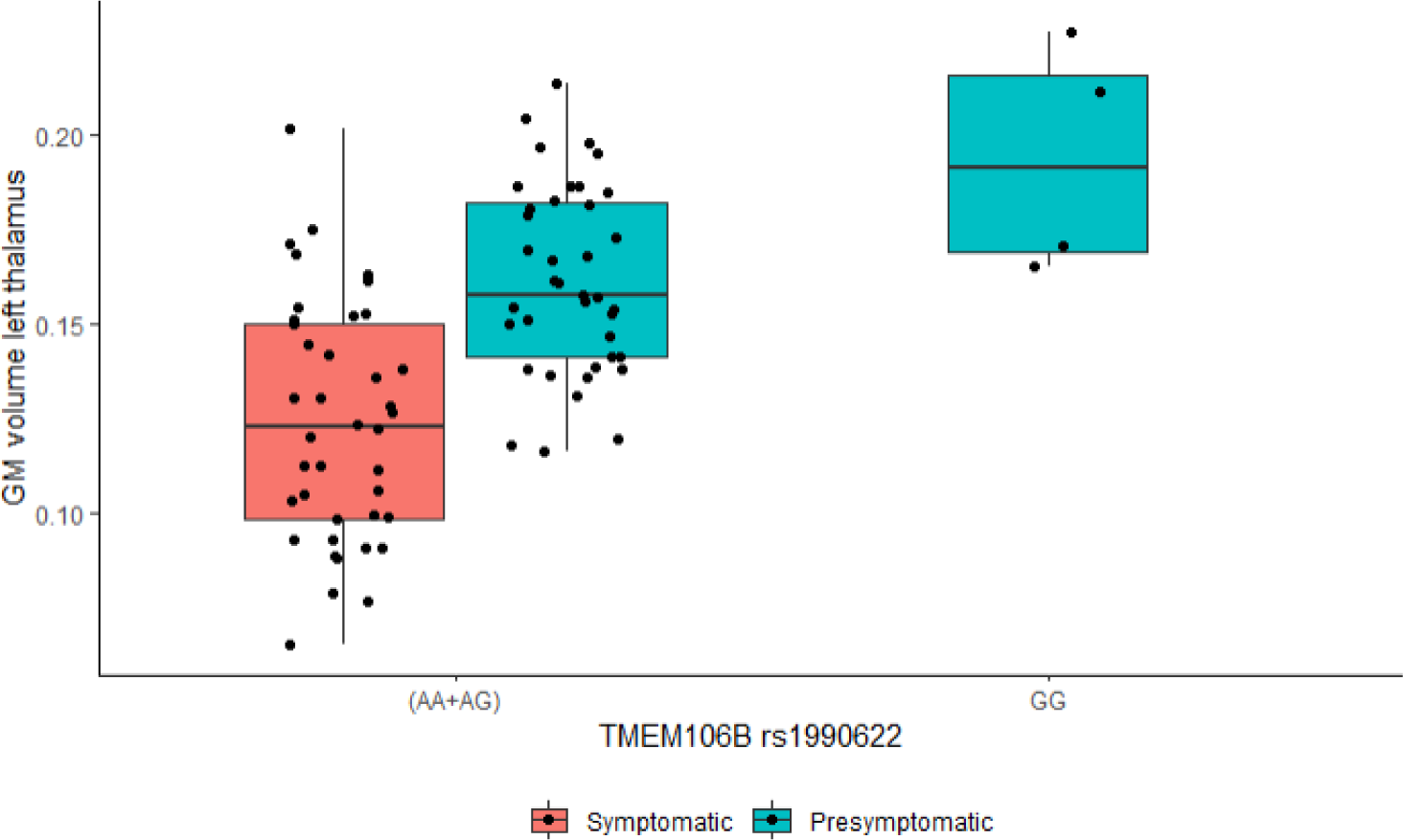
Left thalamic gray matter volume in *GRN* mutation carriers, grouped by symptomatic status and *TMEM106B* rs1990622 genotype dosages.

The mean age of onset of the affected *GRN* mutation carriers in our total cohort with bvFTD, CBS or PPA as primary diagnosis is 59.23 ± 9.23 years old. The presymptomatic *GRN* mutation carriers that carry the *TMEM106B* rs1990622*GG genotype are 29, 45, 49 and 68 years old at their last visit. NfL levels were available for the presymptomatic *GRN* mutation carriers with *TMEM106B* rs1990622*GG with an age at visit of 29 and 68 years old, respectively. **Figure 2** depicts the age at visit and NfL levels for all *GRN* mutation carriers with NfL levels available at the time of imaging. Visually, it can be observed that the presymptomatic *GRN* mutation carrier with *TMEM106B* rs1990622*GG genotype with a current age of 68 years had among the lowest NfL levels (7.967 pg/mL), compared with both symptomatic (mean = 61.250 pg/mL) and presymptomatic *TMEM106B* rs1990622*AA and rs1990622*AG genotype *GRN* mutation carriers (mean = 24.774 pg/mL) within the same age range (65-77 year).

**Figure 2.**
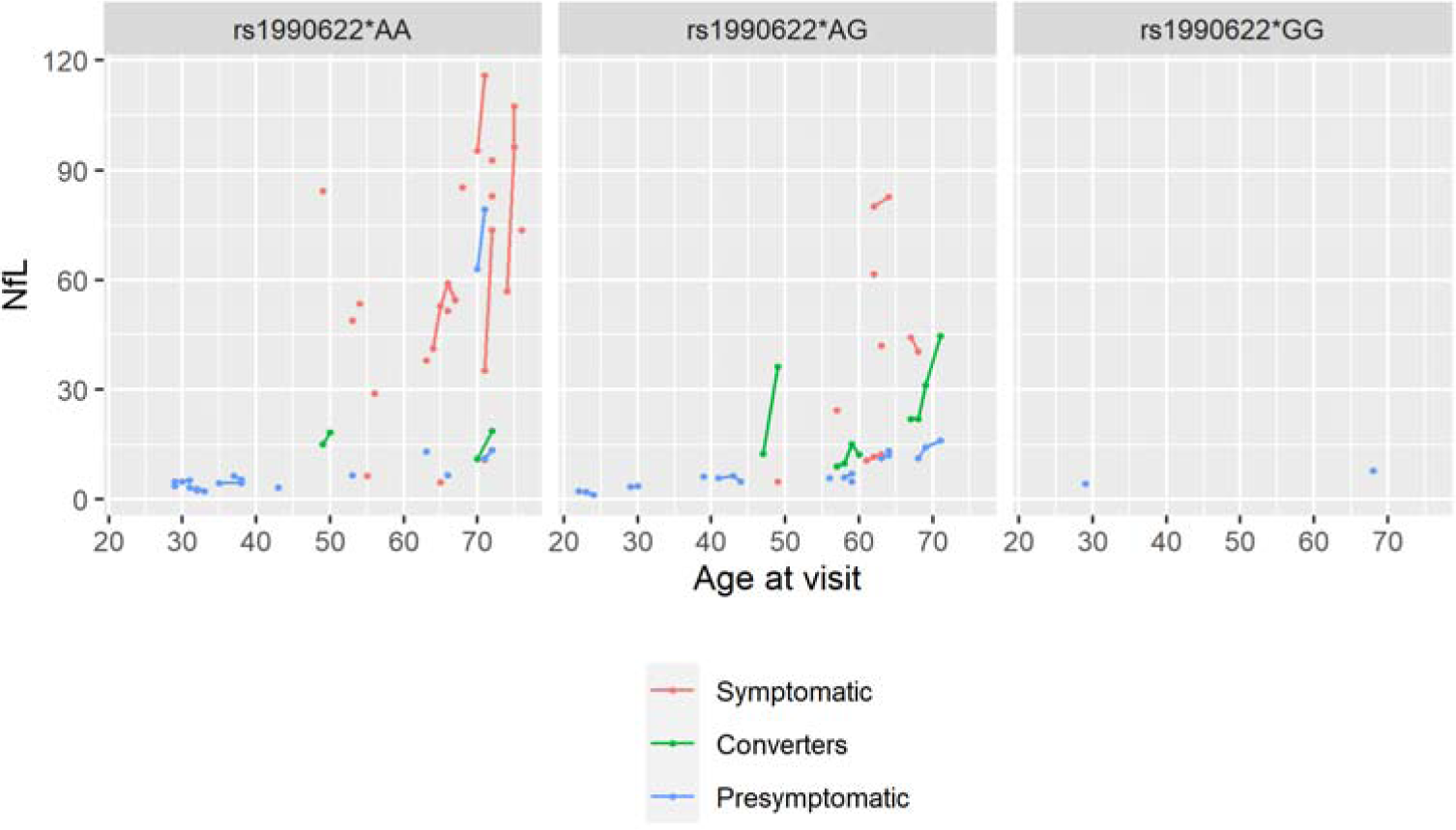
Scatter plot depicting the age at visit (x-axis) and NfL levels (y-axis) for all *GRN* mutation carriers with imaging data and NfL levels measured, according to *TMEM106B* rs1990622 genotype. Blue dots: presymptomatic *GRN* mutation carriers, red dots: symptomatic *GRN* mutation carriers, green dots: *GRN* mutation carriers that converted from presymptomatic to symptomatic status. The lines connect data points that come from the same *GRN* mutation carrier.

Longitudinally, the analyses were conducted with the additive model for *TMEM106B* rs1990622 in affected *GRN* mutation carriers. Statistical analyses were conducted for comparison of the rs1990622*AA group versus rs1990622*AG in affected *GRN* mutation carriers. We found no differences in the rate of decline in total gray matter volume across rs1990622*AG carriers versus rs1990622*AA carriers (beta = 0.536, 95% CI [-1.25, 2.19], *p* = 0.526).

### Association of *TMEM106B* rs1990622 with cognition

In linear mixed models with genetic status, years of education, sex, age at visit and CDR®+ NACC-FTLD sum of boxes score as fixed covariates and pedigree as random effect, *TMEM106B* rs1990622 did not statistically associate with UDS3-EF across the complete cohort, neither in the additive dosage model nor in the recessive model (**eTable 6**), or in subgroup analyses in all affected individuals (**eTable 7**).

Fitting the linear mixed-interaction model between *TMEM106B* rs1990622 and genetic group (none-mutation carrier, *GRN*, *MAPT* or *C9orf72*), with as fixed covariates years of education, sex, age at visit and CDR®+NACC-FTLD sum of boxes and with pedigree as random effect, an effect of *TMEM106B* rs1990622 on UDS3-EF score in *C9orf72* mutation carriers was observed with recessive *TMEM106B* dosages (**Table 4**).

**Table 4.**
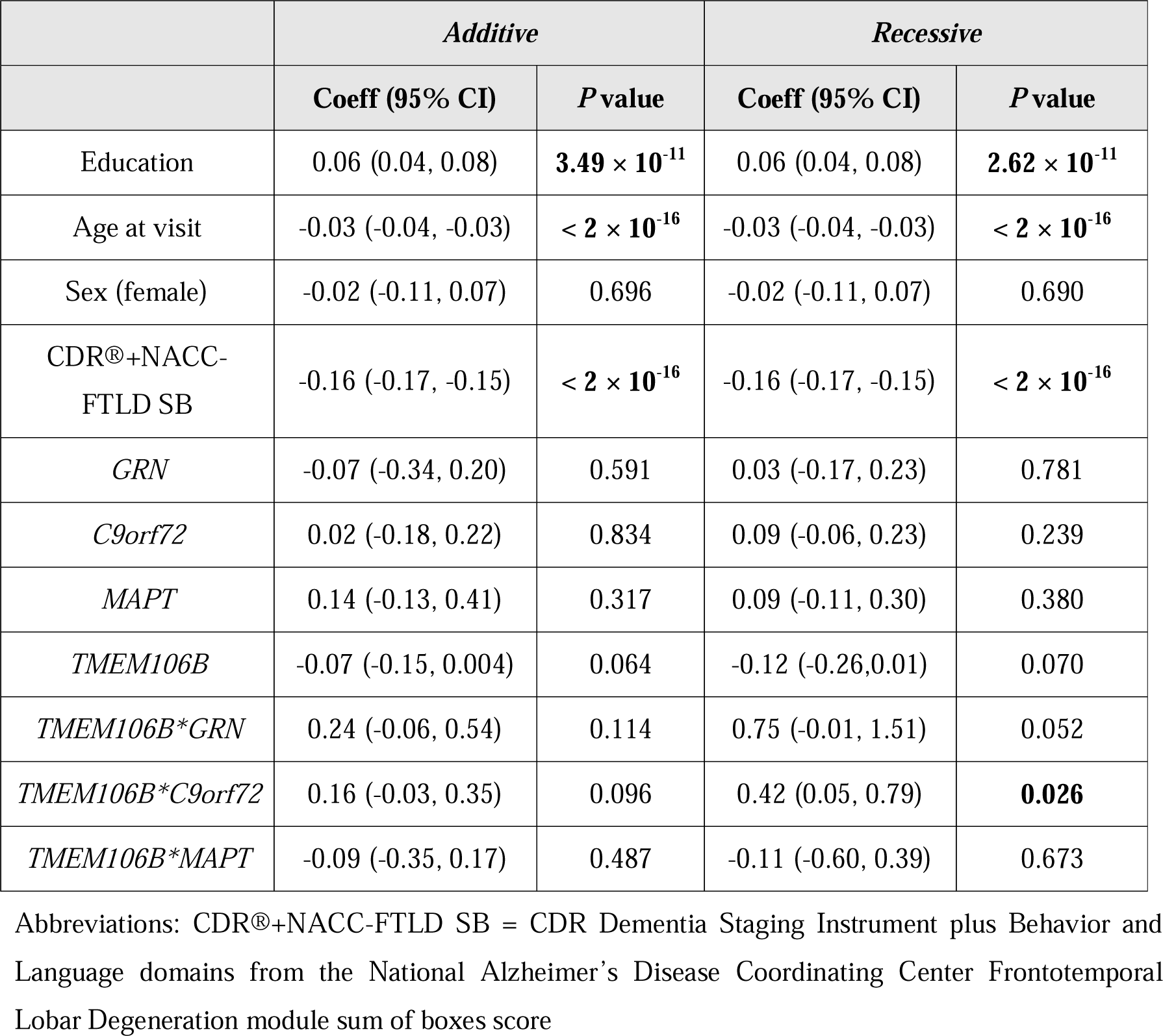
Linear mixed model statistics for *TMEM106B* rs1990622 by genetic group interaction on UDS3-EF.

|  | *Additive* |  | *Recessive* |  |
| --- | --- | --- | --- | --- |
|  | Coeff (95% CI) | *P* value | Coeff (95% CI) | *P* value |
| Education | 0.06 (0.04, 0.08) | $3.49 \times 10^{-11}$ | 0.06 (0.04, 0.08) | $2.62 \times 10^{-11}$ |
| Age at visit | -0.03 (-0.04, -0.03) | $< 2 \times 10^{-16}$ | -0.03 (-0.04, -0.03) | $< 2 \times 10^{-16}$ |
| Sex (female) | -0.02 (-0.11, 0.07) | 0.696 | -0.02 (-0.11, 0.07) | 0.690 |
| CDR®+NACC-FTLD SB | -0.16 (-0.17, -0.15) | $< 2 \times 10^{-16}$ | -0.16 (-0.17, -0.15) | $< 2 \times 10^{-16}$ |
| *GRN* | -0.07 (-0.34, 0.20) | 0.591 | 0.03 (-0.17, 0.23) | 0.781 |
| *C9orf72* | 0.02 (-0.18, 0.22) | 0.834 | 0.09 (-0.06, 0.23) | 0.239 |
| *MAPT* | 0.14 (-0.13, 0.41) | 0.317 | 0.09 (-0.11, 0.30) | 0.380 |
| *TMEM106B* | -0.07 (-0.15, 0.004) | 0.064 | -0.12 (-0.26, 0.01) | 0.070 |
| *TMEM106B\*GRN* | 0.24 (-0.06, 0.54) | 0.114 | 0.75 (-0.01, 1.51) | 0.052 |
| *TMEM106B\*C9orf72* | 0.16 (-0.03, 0.35) | 0.096 | 0.42 (0.05, 0.79) | **0.026** |
| *TMEM106B\*MAPT* | -0.09 (-0.35, 0.17) | 0.487 | -0.11 (-0.60, 0.39) | 0.673 |

Abbreviations: CDR®+NACC-FTLD SB = CDR Dementia Staging Instrument plus Behavior and Language domains from the National Alzheimer's Disease Coordinating Center Frontotemporal Lobar Degeneration module sum of boxes score

In subgroup analyses in *C9orf72* mutation carriers, *TMEM106B* remained associated with UDS3-EF in the recessive model (beta = 0.36, 95% CI [0.05,0.66], *p* = 0.021), and in subgroup analyses in presymptomatic *C9orf72* mutation carriers (beta = 0.33, 95% CI [0.03,0.63], *p* = 0.036). Similar estimates were obtained upon conducting sensitivity analyses in *C9orf72* mutation carriers of European ancestry only (beta = 0.40, 95% CI [0.09, 0.70], *p* = 0.011) and presymptomatic *C9orf72* mutation carriers of European ancestry only (beta = 0.40, 95% CI [0.10, 0.71], *p =* 0.011). In symptomatic *C9orf72* mutation carriers, there was no effect of *TMEM106B* on UDS3-EF (beta = 0.31, 95% CI [-0.19,0.81], *p* = 0.232).

We did not identify statistically significant longitudinal trajectory differences according to *TMEM106B* genotype group (data not shown). In presymptomatic *C9orf72* mutation carriers with at least 2 visits there was no significant decline in cognitive trajectory over time. However, taking into account all the longitudinally collected visits in presymptomatic *C9orf72* mutation carriers, we found in both the additive (beta = 0.22, 95% CI [0.05, 0.39], *p* = 0.014) and recessive (beta = 0.45, 95% CI [0.13, 0.78], *p* = 0.008) model (**eTable 10**), that the minor allele of *TMEM106B* rs1990622 is associated with an increased UDS3-EF score, in line with the cross-sectional data taking only the last visit into account.

## DISCUSSION

*TMEM106B* was initially identified as genetic risk factor for FTLD-TDP. Since then, it has been shown to not only act as modifier of disease penetrance in FTLD-TDP but also as modifier of pathological, imaging and clinical characteristics of FTD and related neurodegenerative diseases. To further confirm the association of *TMEM106B* SNPs with imaging and clinical characteristics in FTD and to evaluate its role in the different genetic groups of autosomal dominant FTD we performed association analyses in the largest available systematically ascertained cohort of FTD patients.

In our complete cohort with imaging data available, no significant association of gray matter brain volumes with *TMEM106B* were detected. However, in *GRN* mutation carriers, carrying two copies of the minor allele of *TMEM106B* was associated with a larger total gray matter volume. This was most pronounced in the thalamus in the left hemisphere, a finding that remained in a subgroup of presymptomatic *GRN* mutation carriers. Thalamic atrophy is a common feature in frontotemporal dementia, and especially in *GRN* mutation carriers, asymmetry in thalamic volumes is apparent^36^. Furthermore, *GRN* presymptomatic mutation carriers display changes in intrinsic connectivity networks, with the thalamus as key hub^37^. This is in line with findings in mice with homozygous *GRN* deletions (*GRN*^-/-^)^38^, where microglial activation in the ventral thalamus drives neurodegeneration in the thalamocortical circuit^38^. Interestingly, patients with FTLD-*GRN* and *GRN*^-/-^ mice show similar transcriptomic and histopathological changes in the thalamus, not only in microglia but also in astrocytes, promoting neurodegeneration^39^. Other regions that appear altered in response to *TMEM106B* are the frontal, temporal, parietal, anterior cingulate areas, insula and cerebellum, in line with known patterns of atrophy described in *GRN* mutation carriers^40–42^ and in patients with FTLD-TDP type A, the pathology uniformly present in patients with *GRN* mutations. In addition, previous research showed an effect of *TMEM106B* in these regions in a clinically diagnosed FTD cohort^16^.

Importantly, the *GRN* mutation carriers with two copies of the minor allele of *TMEM106B* were all presymptomatic at time of imaging. With a mean age of onset of 59 years in affected *GRN* mutation carriers in our total cohort, it cannot be excluded that these presymptomatic *GRN* mutation carriers will still develop FTD at a later age; however, one of these presymptomatic *GRN* mutation carriers remained without symptoms at 68 years of age. The strikingly low NfL level of this participant compared to *GRN* mutation carriers within the same age range (65-76 years), also well below the mean value of phenoconverters^43^, supports the hypothesis that carrying two copies of the minor allele of *TMEM106B* might offer protection against developing FTD, or at a minimum a delay in disease onset.

In *C9orf72* we did not observe an association between *TMEM106B* and (sub)cortical atrophy. In fact, at the presymptomatic stage, we found that irrespective of the *TMEM106B* genotype, the presence of *C9orf72* is associated with lower gray matter volumes in comparison to clinically normal non-mutation carriers, consistent with prior work showing structural brain changes occurring 10 to 40 years before onset^27,40,44^. In *GRN* mutation carriers, on the other hand, changes in brain volume occur only within a few years proximity to onset of symptomatic FTD^27,41,42,44^. Moreover, while the rate of volume loss differs between *C9orf72* and *GRN,* with attenuated atrophy rate after onset of symptomatic FTD in *C9orf72* and with an acceleration of atrophy rate after onset in *GRN,* their rate of functional decline is similar^42^. Hence, there might be earlier and divergent pathophysiological changes in *C9orf72* as compared to *GRN* mutation carriers in the presymptomatic phase, with the early loss of gray matter volume in *C9orf72* mutation carriers masking a potential effect of *TMEM106B*.

Contrary to structural imaging, we did identify a protective effect of the *TMEM106B* rs1990622 minor allele on cognition in *C9orf72*, especially in presymptomatic *C9orf72* mutation carriers. With participants with a *C9orf72* repeat expansion already showing signs of neurodegeneration (e.g. gray matter loss) prior to symptom onset, we hypothesize that *TMEM106B* is able to modulate the resilience against developing clinical FTD during these early stages of disease. In support of this hypothesis, homozygosity for the minor allele has been shown to protect *C9orf72* carriers from developing FTD but not from developing ALS^11^. Moreover, discordance between the presence of disease pathology and effects on cognition in the aging population is a known phenomenon and *TMEM106B* has been suggested as a potential modifier of this ‘cognitive resilience’, with the minor allele of *TMEM106B* rs1990622 being associated with a better performance than expected based on pathological burden^45^.

Previous studies focusing on presymptomatic genetic FTD have identified modulating effects of *TMEM106B* genotype on gray matter volume in mutation carriers (combining *GRN, C9orf72, MAPT*) versus non-carrier family controls^17,18^. Importantly, a different distribution in genetic groups between our study and the previously conducted studies^17,18^ can be noted, with *GRN* being the largest group and *MAPT* being the smallest group in the previous studies while in this study the mutation carriers are enriched for *C9orf72* and *MAPT* carriers, with *GRN* being the smallest group (22% versus 56% in the previous studies). Furthermore, in this study we also included a sporadic FTD cohort without mutations identified in the known FTD genes. Hence, we investigated the association of *TMEM106B* with gray matter volume and cognition in each genetic group separately via interaction modelling and subgroup analyses. We identified associations of *TMEM106B* in the *GRN* and *C9orf72* genetic groups. This is in line with *TMEM106B* being identified as modifier in those with TDP-43 pathology^5,6^ but not in most other clinical FTD cohorts of non-TDP^46^ or unknown pathology^5,47,48^, with a few exceptions^7,16^ potentially due to a substantial proportion of cases with FTLD-TDP pathology^7,16^. Beyond FTLD-TDP, *TMEM106B* is associated with hippocampal sclerosis of aging^21,49,50^, with or without accompanying Alzheimer type pathology, with hippocampal sclerosis in Lewy body disease^51^, and with limbic-predominant age-related TDP-43 proteinopathy (LATE-NC)^52,53^, all characterized by the presence of TDP-43 proteinopathy. Furthermore, TDP-43 inclusions are also present in Alzheimer’s disease and Parkinson’s disease^54^, explaining the broader modifying roles of *TMEM106B* on endophenotypes such as cognition across neurodegenerative diseases.

Strikingly, TMEM106B filaments form aggregates in the brain in elderly and across neurodegenerative diseases^55,56^, with the risk allele associated with greater fibril formation^57^ and enhanced TDP-43 dysfunction^58^. While fibril accumulation has been found to be a common age-related phenomenon, fibril aggregates were especially abundant in patients with *GRN* mutations^59^. Both progranulin and TMEM106B, are important players in lysosomal health^55^. TMEM106B is a transmembrane glycoprotein that primarily localizes to lysosomal membranes where it is proteolytically processed. Progranulin is cleaved in the lysosome into functional granulins, and homozygous loss-of-function mutations in *GRN* cause the lysosomal storage disorder neuronal ceroid lipofuscinosis 11. In addition to convergence of pathomechanisms between *GRN* and *TMEM106B*, TMEM106B-induced lysosomal defects due to increased TMEM106B expression have been shown to be C9orf72-dependent^60^. Altogether, these studies support a specific role for *TMEM106B* as modifier in FTLD-TDP pathophysiology.

We acknowledge that there are limitations with this work. Although we investigated modifying effects of *TMEM106B* in the largest collection of systematically ascertained FTD patients and families from the ALLFTD study, the number of individuals with a *GRN* mutation and two copies of the minor (protective) allele of *TMEM106B*, are small. While this supports a role for *TMEM106B* in reducing disease penetrance, extensive recruitment of unaffected family members of *GRN* mutation carriers followed by genetic analyses of *TMEM106B* and *GRN* will be required to specifically identify those individuals that carry a *GRN* mutation and two copies of the *TMEM106B* minor allele. In addition, to reach the maximum sample size for each outcome measure of interest, the last visit with the measure of interest available was selected. In this way, the analyses differ in their set of unique individuals and their respective time point of assessment, precluding multivariate analysis of variance studies to assess simultaneously associations between *TMEM106B,* imaging and cognition in the same cohort. While we used the largest data set possible, some of our negative statistical associations may be due to small sample sizes. Despite these limitations, we confirmed *TMEM106B* as modifier in *GRN* and *C9orf72* mutation carriers, and reported distinct effects in different genetic groups. Importantly, we showed that *TMEM106B* already exerts effects in the presymptomatic stage of disease. With clinical trials ongoing for gene-based therapies for *GRN* and *C9orf72* mutation carriers, it is important to take *TMEM106B* genetic status into account in the clinical trial design and recruitment of participants.

## ACKNOWLEDGEMENTS

We wish to express our gratitude towards all ALLFTD participants and their caregivers for the dedicated participation in this research program. We also thank all personnel involved in the ALLFTD consortium who are not listed as co-authors.

## REFERENCES

1. Baker M, Mackenzie IR, Pickering-Brown SM, et al. Mutations in progranulin cause tau-negative frontotemporal dementia linked to chromosome 17. Nature. Aug 24 2006;442(7105):916-9. doi:10.1038/nature05016

2. Cruts M, Gijselinck I, van der Zee J, et al. Null mutations in progranulin cause ubiquitin-positive frontotemporal dementia linked to chromosome 17q21. Nature. Aug 24 2006;442(7105):920–4. doi:10.1038/nature05017

3. Hutton M, Lendon CL, Rizzu P, et al. Association of missense and 5’-splice-site mutations in tau with the inherited dementia FTDP-17. Nature. Jun 18 1998;393(6686):702–5. doi:10.1038/31508

4. DeJesus-Hernandez M, Mackenzie IR, Boeve BF, et al. Expanded GGGGCC hexanucleotide repeat in noncoding region of C9ORF72 causes chromosome 9p-linked FTD and ALS. Neuron. Oct 20 2011;72(2):245–56. doi:10.1016/j.neuron.2011.09.011

5. Van Deerlin VM, Sleiman PM, Martinez-Lage M, et al. Common variants at 7p21 are associated with frontotemporal lobar degeneration with TDP-43 inclusions. Nat Genet. Mar 2010;42(3):234–9. doi:10.1038/ng.536

6. Pottier C, Zhou X, Perkerson RB, 3rd, et al. Potential genetic modifiers of disease risk and age at onset in patients with frontotemporal lobar degeneration and GRN mutations: a genome-wide association study. Lancet Neurol. Jun 2018;17(6):548–558. doi:10.1016/S1474-4422(18)30126-1

7. van der Zee J, Van Langenhove T, Kleinberger G, et al. TMEM106B is associated with frontotemporal lobar degeneration in a clinically diagnosed patient cohort. Brain. Mar 2011;134(Pt 3):808–15. doi:10.1093/brain/awr007

8. Finch N, Carrasquillo MM, Baker M, et al. TMEM106B regulates progranulin levels and the penetrance of FTLD in GRN mutation carriers. Neurology. Feb 1 2011;76(5):467–74. doi:10.1212/WNL.0b013e31820a0e3b

9. Lattante S, Le Ber I, Galimberti D, et al. Defining the association of TMEM106B variants among frontotemporal lobar degeneration patients with GRN mutations and C9orf72 repeat expansions. Neurobiol Aging. Nov 2014;35(11):2658 e1-2658 e5. doi:10.1016/j.neurobiolaging.2014.06.023

10. Perneel J, Manoochehri M, Huey ED, Rademakers R, Goldman J. Case report: TMEM106B haplotype alters penetrance of GRN mutation in frontotemporal dementia family. Front Neurol. 2023;14:1160248. doi:10.3389/fneur.2023.1160248

11. van Blitterswijk M, Mullen B, Nicholson AM, et al. TMEM106B protects C9ORF72 expansion carriers against frontotemporal dementia. Acta Neuropathol. Mar 2014;127(3):397–406. doi:10.1007/s00401-013-1240-4

12. Adams HH, Verhaaren BF, Vrooman HA, et al. TMEM106B influences volume of left-sided temporal lobe and interhemispheric structures in the general population. Biol Psychiatry. Sep 15 2014;76(6):503–8. doi:10.1016/j.biopsych.2014.03.006

13. Yu L, De Jager PL, Yang J, Trojanowski JQ, Bennett DA, Schneider JA. The TMEM106B locus and TDP-43 pathology in older persons without FTLD. Neurology. Mar 3 2015;84(9):927–34. doi:10.1212/WNL.0000000000001313

14. Rhinn H, Abeliovich A. Differential Aging Analysis in Human Cerebral Cortex Identifies Variants in TMEM106B and GRN that Regulate Aging Phenotypes. Cell Syst. Apr 26 2017;4(4):404–415 e5. doi:10.1016/j.cels.2017.02.009

15. Li Z, Farias FHG, Dube U, et al. The TMEM106B FTLD-protective variant, rs1990621, is also associated with increased neuronal proportion. Acta Neuropathol. Jan 2020;139(1):45–61. doi:10.1007/s00401-019-02066-0

16. Harding SR, Bocchetta M, Gordon E, et al. The TMEM106B risk allele is associated with lower cortical volumes in a clinically diagnosed frontotemporal dementia cohort. J Neurol Neurosur Ps. Nov 2017;88(11):997-+. doi:10.1136/jnnp-2017-315641

17. Gazzina S, Grassi M, Premi E, et al. Education modulates brain maintenance in presymptomatic frontotemporal dementia. J Neurol Neurosurg Psychiatry. Oct 2019;90(10):1124–1130. doi:10.1136/jnnp-2019-320439

18. Premi E, Grassi M, van Swieten J, et al. Cognitive reserve and TMEM106B genotype modulate brain damage in presymptomatic frontotemporal dementia: a GENFI study. Brain. Jun 1 2017;140(6):1784–1791. doi:10.1093/brain/awx103

19. Premi E, Formenti A, Gazzina S, et al. Effect of TMEM106B Polymorphism on Functional Network Connectivity in Asymptomatic GRN Mutation Carriers. Jama Neurol. Feb 2014;71(2):216–221. doi:10.1001/jamaneurol.2013.4835

20. Tropea TF, Mak J, Guo MH, et al. TMEM106B Effect on cognition in Parkinson disease and frontotemporal dementia. Annals of Neurology. Jun 2019;85(6):801–811. doi:10.1002/ana.25486

21. Rutherford NJ, Carrasquillo MM, Li M, et al. TMEM106B risk variant is implicated in the pathologic presentation of Alzheimer disease. Neurology. Aug 14 2012;79(7):717–8. doi:10.1212/WNL.0b013e318264e3ac

22. Vass R, Ashbridge E, Geser F, et al. Risk genotypes at TMEM106B are associated with cognitive impairment in amyotrophic lateral sclerosis. Acta Neuropathol. Mar 2011;121(3):373–80. doi:10.1007/s00401-010-0782-y

23. Manini A, Ratti A, Brusati A, et al. TMEM106B Acts as a Modifier of Cognitive and Motor Functions in Amyotrophic Lateral Sclerosis. Int J Mol Sci. Aug 2022;23(16) doi: ARTN 9276 10.3390/ijms23169276

24. Boeve B, Bove J, Brannelly P, et al. The longitudinal evaluation of familial frontotemporal dementia subjects protocol: Framework and methodology. Alzheimers Dement. Jan 2020;16(1):22–36. doi:10.1016/j.jalz.2019.06.4947

25. Ramos EM, Dokuru DR, Van Berlo V, et al. Genetic screening of a large series of North American sporadic and familial frontotemporal dementia cases. Alzheimers Dement. Jan 2020;16(1):118–130. doi:10.1002/alz.12011

26. Ramos EM, Wojta K, Yang Z, et al. Inferring genetic relatedness in a large, multisite frontotemporal dementia series: Data from the ALLFTD consortium. Alzheimer’s & Dementia. 2023/06/01 2023;19(S1):e068040. 10.1002/alz.068040

27. Staffaroni AM, Quintana M, Wendelberger B, et al. Temporal order of clinical and biomarker changes in familial frontotemporal dementia. Nat Med. Oct 2022;28(10):2194–2206. doi:10.1038/s41591-022-01942-9

28. Sled JG, Zijdenbos AP, Evans AC. A nonparametric method for automatic correction of intensity nonuniformity in MRI data. IEEE Trans Med Imaging. Feb 1998;17(1):87–97. doi:10.1109/42.668698

29. Ashburner J, Friston KJ. Unified segmentation. Neuroimage. Jul 1 2005;26(3):839-51. doi:10.1016/j.neuroimage.2005.02.018

30. Ashburner J, Friston KJ. Diffeomorphic registration using geodesic shooting and Gauss-Newton optimisation. Neuroimage. Apr 1 2011;55(3):954–67. doi:10.1016/j.neuroimage.2010.12.049

31. Desikan RS, Segonne F, Fischl B, et al. An automated labeling system for subdividing the human cerebral cortex on MRI scans into gyral based regions of interest. Neuroimage. Jul 1 2006;31(3):968–80. doi:10.1016/j.neuroimage.2006.01.021

32. Staffaroni AM, Asken BM, Casaletto KB, et al. Development and validation of the Uniform Data Set (v3.0) executive function composite score (UDS3-EF). Alzheimers Dement. Apr 2021;17(4):574–583. doi:10.1002/alz.12214

33. Asken BM, Ljubenkov PA, Staffaroni AM, et al. Plasma inflammation for predicting phenotypic conversion and clinical progression of autosomal dominant frontotemporal lobar degeneration. J Neurol Neurosurg Psychiatry. Jul 2023;94(7):541–549. doi:10.1136/jnnp-2022-330866

34. Casaletto KB, Kornack J, Paolillo EW, et al. Association of Physical Activity With Neurofilament Light Chain Trajectories in Autosomal Dominant Frontotemporal Lobar Degeneration Variant Carriers. Jama Neurol. Jan 1 2023;80(1):82–90. doi:10.1001/jamaneurol.2022.4178

35. Gendron TF, Heckman MG, White LJ, et al. Comprehensive cross-sectional and longitudinal analyses of plasma neurofilament light across FTD spectrum disorders. Cell Rep Med. Apr 19 2022;3(4):100607. doi:10.1016/j.xcrm.2022.100607

36. Bocchetta M, Gordon E, Cardoso MJ, et al. Thalamic atrophy in frontotemporal dementia - Not just a C9orf72 problem. Neuroimage Clin. 2018;18:675–681. doi:10.1016/j.nicl.2018.02.019

37. Lee SE, Sias AC, Kosik EL, et al. Thalamo-cortical network hyperconnectivity in preclinical progranulin mutation carriers. Neuroimage Clin. 2019;22:101751. doi:10.1016/j.nicl.2019.101751

38. Lui H, Zhang J, Makinson SR, et al. Progranulin Deficiency Promotes Circuit-Specific Synaptic Pruning by Microglia via Complement Activation. Cell. May 5 2016;165(4):921–35. doi:10.1016/j.cell.2016.04.001

39. Marsan E, Velmeshev D, Ramsey A, et al. Astroglial toxicity promotes synaptic degeneration in the thalamocortical circuit in frontotemporal dementia with GRN mutations. J Clin Invest. Mar 15 2023;133(6)doi:10.1172/JCI164919

40. Rohrer JD, Nicholas JM, Cash DM, et al. Presymptomatic cognitive and neuroanatomical changes in genetic frontotemporal dementia in the Genetic Frontotemporal dementia Initiative (GENFI) study: a cross-sectional analysis. Lancet Neurol. Mar 2015;14(3):253–62. doi:10.1016/S1474-4422(14)70324-2

41. Whitwell JL, Boeve BF, Weigand SD, et al. Brain atrophy over time in genetic and sporadic frontotemporal dementia: a study of 198 serial magnetic resonance images. Eur J Neurol. May 2015;22(5):745–52. doi:10.1111/ene.12675

42. Staffaroni AM, Goh SM, Cobigo Y, et al. Rates of Brain Atrophy Across Disease Stages in Familial Frontotemporal Dementia Associated With MAPT, GRN, and C9orf72 Pathogenic Variants. JAMA Netw Open. Oct 1 2020;3(10):e2022847. doi:10.1001/jamanetworkopen.2020.22847

43. Rojas JC, Wang P, Staffaroni AM, et al. Plasma Neurofilament Light for Prediction of Disease Progression in Familial Frontotemporal Lobar Degeneration. Neurology. May 4 2021;96(18):e2296–e2312. doi:10.1212/WNL.0000000000011848

44. Finger E, Malik R, Bocchetta M, et al. Neurodevelopmental effects of genetic frontotemporal dementia in young adult mutation carriers. Brain. May 2 2023;146(5):2120–2131. doi:10.1093/brain/awac446

45. White CC, Yang HS, Yu L, et al. Identification of genes associated with dissociation of cognitive performance and neuropathological burden: Multistep analysis of genetic, epigenetic, and transcriptional data. PLoS Med. Apr 2017;14(4):e1002287. doi:10.1371/journal.pmed.1002287

46. Rostgaard N, Roos P, Budtz-Jorgensen E, et al. TMEM106B and ApoE polymorphisms in CHMP2B-mediated frontotemporal dementia (FTD-3). Neurobiol Aging. Nov 2017;59:221 e1–221 e7. doi:10.1016/j.neurobiolaging.2017.06.026

47. Rollinson S, Mead S, Snowden J, et al. Frontotemporal lobar degeneration genome wide association study replication confirms a risk locus shared with amyotrophic lateral sclerosis. Neurobiol Aging. Apr 2011;32(4):758 e1-7. doi:10.1016/j.neurobiolaging.2010.12.005

48. Nicholson AM, Rademakers R. What we know about TMEM106B in neurodegeneration. Acta Neuropathol. Nov 2016;132(5):639–651. doi:10.1007/s00401-016-1610-9

49. Nelson PT, Wang WX, Partch AB, et al. Reassessment of risk genotypes (GRN, TMEM106B, and ABCC9 variants) associated with hippocampal sclerosis of aging pathology. J Neuropathol Exp Neurol. Jan 2015;74(1):75–84. doi:10.1097/NEN.0000000000000151

50. Murray ME, Cannon A, Graff-Radford NR, et al. Differential clinicopathologic and genetic features of late-onset amnestic dementias. Acta Neuropathol. Sep 2014;128(3):411–21. doi:10.1007/s00401-014-1302-2

51. Aoki N, Murray ME, Ogaki K, et al. Hippocampal sclerosis in Lewy body disease is a TDP-43 proteinopathy similar to FTLD-TDP Type A. Acta Neuropathol. Jan 2015;129(1):53–64. doi:10.1007/s00401-014-1358-z

52. Neumann M, Perneel J, Cheung S, et al. Limbic-predominant age-related TDP-43 proteinopathy (LATE-NC) is associated with abundant TMEM106B pathology. Acta Neuropathol. Jul 2023;146(1):163–166. doi:10.1007/s00401-023-02580-2

53. Dugan AJ, Nelson PT, Katsumata Y, et al. Analysis of genes (TMEM106B, GRN, ABCC9, KCNMB2, and APOE) implicated in risk for LATE-NC and hippocampal sclerosis provides pathogenetic insights: a retrospective genetic association study. Acta Neuropathol Commun. Sep 15 2021;9(1):152. doi:10.1186/s40478-021-01250-2

54. Cook C, Zhang YJ, Xu YF, Dickson DW, Petrucelli L. TDP-43 in neurodegenerative disorders. Expert Opin Biol Ther. Jul 2008;8(7):969–78. doi:10.1517/14712598.8.7.969

55. Perneel J, Rademakers R. Identification of TMEM106B amyloid fibrils provides an updated view of TMEM106B biology in health and disease. Acta Neuropathol. Nov 2022;144(5):807–819. doi:10.1007/s00401-022-02486-5

56. Chang A, Xiang X, Wang J, et al. Homotypic fibrillization of TMEM106B across diverse neurodegenerative diseases. Cell. Apr 14 2022;185(8):1346–1355 e15. doi:10.1016/j.cell.2022.02.026

57. Lee JY, Harney DJ, Teo JD, et al. The major TMEM106B dementia risk allele affects TMEM106B protein levels, fibril formation, and myelin lipid homeostasis in the ageing human hippocampus. Mol Neurodegener. Sep 19 2023;18(1):63. doi:10.1186/s13024-023-00650-3

58. Marks JD, Ayuso VE, Carlomagno Y, et al. TMEM106B core deposition associates with TDP-43 pathology and is increased in risk SNP carriers for frontotemporal dementia. Sci Transl Med. Jan 17 2024;16(730):eadf9735. doi:10.1126/scitranslmed.adf9735

59. Vicente CT, Perneel J, Wynants S, et al. C-terminal TMEM106B fragments in human brain correlate with disease-associated TMEM106B haplotypes. Brain. Oct 3 2023;146(10):4055–4064. doi:10.1093/brain/awad133

60. Busch JI, Unger TL, Jain N, Tyler Skrinak R, Charan RA, Chen-Plotkin AS. Increased expression of the frontotemporal dementia risk factor TMEM106B causes C9orf72-dependent alterations in lysosomes. Hum Mol Genet. Jul 1 2016;25(13):2681–2697. doi:10.1093/hmg/ddw127

