## Supplementary Material for "Gene specific effects on brain volume and cognition of *TMEM106B* in frontotemporal lobar degeneration"

*Corresponding author:

VIB Center for Molecular Neurology

**eTable 1. Individual regions of interest (ROIs) for gray matter volumes. Left and right hemispheres were considered separately in analyses**

| Cortical volumes | Subcortical volumes | Cerebellar volume |
|---|---|---|
| Frontal lobe | Hippocampus | |
| Temporal lobe | Amygdala | |
| Parietal lobe | Caudate | |
| Occipital lobe | Accumbens | Cerebellum |
| Cingulate cortices: rostral anterior, caudal anterior, isthmus, posterior | Pallidum | |
| Insular cortices | Thalamus | |
| | Putamen | |

**eTable 2. Demographic characteristics for participants included in the analyses with gray matter volume as outcome (N = 958)**

| Characteristic | All mutation carriers | C9orf72+ | GRN+ | MAPT+ | Non-carriers |
|---|---|---|---|---|---|
| Sample size | 373 | 169 | 82 | 106 | 585 |
| Age at visit (yr), mean (s.d) | 51.19 (14.14) | 51.18 (13.97) | 57.63 (13.25) | 46.7 (12.57) | 58.59 (13.79) |
| Female, n (%) | 203 (54.42 %) | 92 (54.44 %) | 40 (48.78 %) | 62 (58.49 %) | 299 (51.11 %) |
| Education (yr), mean (s.d) | 15.67 (2.48) | 15.75 (2.39) | 15.72 (2.75) | 15.54 (2.48) | 15.95 (2.50) |
| **Race, n** | | | | | |
| EUR | 361 | 167 | 79 | 101 | 544 |
| Non-EUR | 11 | 1 | 3 | 5 | 34 |
| Unknown | 1 | 1 | 0 | 0 | 7 |
| ***TMEM106B* rs1990622, n** | | | | | |
| A/A | 161 | 71 | 42 | 42 | 204 |
| A/G | 161 | 71 | 36 | 50 | 277 |
| G/G | 51 | 27 | 4 | 14 | 104 |
| **CDR®+NACC-FTLD Global, n** | | | | | |
| 0 | 191 | 92 | 44 | 47 | 235 |
| 0.5 | 71 | 38 | 11 | 19 | 83 |

| | | | | | |
|---|---|---|---|---|---|
| ≥ 1 | 111 | 39 | 27 | 40 | 267 |
| **CDR®+NACC-FTLD sum of boxes, mean (s.d)** | 3.22 (5.11) | 2.52 (4.40) | 3.82 (5.65) | 3.93 (5.70) | 4.03 (4.88) |
| **Primary clinical phenotype, n** | | | | | |
| Clinically normal | 188 | 91 | 42 | 48 | 234 |
| MBI/MCI | 41 | 19 | 8 | 14 | 32 |
| bvFTD | 91 | 35 | 15 | 38 | 122 |
| ALS | 6 | 6 | 0 | 0 | 0 |
| FTD-ALS | 5 | 4 | 0 | 0 | 5 |
| PPA | o | 3 | 9 | 0 | 87 |
| CBS | 3 | 0 | 2 | 1 | 49 |
| PSP | 3 | 0 | 1 | 2 | 56 |
| Other | 21 | 11 | 5 | 3 | 0 |
| **MoCA, mean (s.d.)** | 24.42 (6.29)<br>NA: 31 | 25.29 (5.22)<br>NA: 15 | 22.8 (7.87)<br>NA: 7 | 24.09 (6.71)<br>NA: 7 | 22.81 (6.91)<br>NA: 54 |
| **Total gray matter volume (% of total intracranial volume) mean (s.d)** | 41.91 (5.62) | 41.69 (5.36) | 40.68 (5.79) | 42.87 (5.66) | 41.70 (6.91) |

**eTable 3. Demographic characteristics for participants included in the analyses with UDS3-EF as outcome (N =1581)**

| Characteristic | All mutation carriers | C9orf72+ | GRN+ | MAPT+ | Non-carriers |
|---|---|---|---|---|---|
| Sample size | 465 | 229 | 101 | 113 | 1116 |
| Age at visit (yr), mean (s.d) | 53.17 (14.24) | 53.05 (14.19) | 58.39 (12.97) | 48.44 (12.87) | 62.1 (12.47) |
| Female, n (%) | 248 (53.33 %) | 126 (55.02 %) | 46 (45.54 %) | 65 (57.52 %) | 527 (47.22 %) |
| Education (yr), mean (s.d) | 15.55 (2.51) | 15.62 (2.39) | 15.5 (2.89) | 15.62 (2.42) | 16.06 (2.61) |
| **Race, n** | | | | | |
| EUR | 448 | 225 | 95 | 108 | 1021 |
| Non-EUR | 13 | 1 | 5 | 5 | 79 |
| Unknown | 4 | 3 | 1 | 0 | 16 |
| ***TMEM106B* rs1990622, n** | | | | | |
| A/A | 192 | 90 | 49 | 44 | 348 |
| A/G | 212 | 108 | 46 | 53 | 551 |
| G/G | 61 | 31 | 6 | 16 | 217 |
| **CDR®+NACC-FTLD Global, n** | | | | | |
| 0 | 208 | 108 | 43 | 47 | 279 |
| 0.5 | 78 | 41 | 14 | 20 | 181 |

| | | | | | |
|---|---|---|---|---|---|
| ≥ 1 | 179 | 80 | 44 | 46 | 656 |
| **CDR®+NACC-FTLD sum of boxes, mean (s.d)** | 4.08 (5.35) | 3.78 (5.33) | 4.93 (5.96) | 3.89 (4.71) | 5.27 (4.90) |
| **Primary clinical phenotype, n** | | | | | |
| Clinically normal | 202 | 106 | 42 | 46 | 277 |
| MBI/MCI | 46 | 22 | 9 | 14 | 56 |
| bvFTD | 144 | 65 | 28 | 44 | 285 |
| ALS | 10 | 10 | 0 | 0 | 0 |
| FTD-ALS | 11 | 10 | 0 | 0 | 19 |
| PPA | 15 | 3 | 8 | 1 | 204 |
| CBS | 9 | 1 | 6 | 1 | 108 |
| PSP | 4 | 2 | 1 | 1 | 167 |
| Other | 24 | 10 | 7 | 6 | 0 |
| **MoCA, mean (s.d.)** | 22.66 (7.49)<br>NA: 21 | 23.39 (6.93)<br>NA: 13 | 20.8 (8.42)<br>NA: 4 | 22.76 (7.53)<br>NA: 4 | 21.09 (7.18)<br>NA: 60 |
| **USD3-EF (composite z-score) mean (s.d)** | -0.65 (1.49) | -0.59 (1.42) | -0.97 (1.52) | -0.49 (1.60) | -1.29 (1.38) |

**eTable 4. Number of individuals grouped by gene-affection status, included in the fitted linear mixed models to investigate the association between the gene-affection status on the outcome measures**

| Group | Description | GM volume N=942 | UDS3-EF N = 1559 | ALSFRS-R N = 1003 | UPDRS N = 1087 |
|---|---|---|---|---|---|
| 1 | Non-carrier & asymptomatic | 234 | 277 | / | / |
| 2 | Non-carrier & symptomatic | 351 | 839 | 746 | 838 |
| 3 | GRN & asymptomatic | 42 | 42 | / | / |
| 4 | GRN & symptomatic | 40 | 59 | 64 | 59 |
| 5 | C9orf72 & asymptomatic | 91 | 106 | / | / |
| 6 | C9orf72 & symptomatic | 78 | 123 | 121 | 119 |
| 7 | MAPT & asymptomatic | 48 | 46 | / | / |
| 8 | MAPT & symptomatic | 58 | 67 | 72 | 71 |

**eTable 5. Linear mixed models were fitted to investigate the association between the gene-affection status on the gray matter and cognitive (UDS3-EF) outcome measures, adjusted for education, age, sex and CDR+NACC-FTLD sum of boxes score as fixed effects and pedigree as random effect.**

| | ***Total gray matter volume N = 942*** | | ***UDS3-EF N = 1559*** | |
|---|---|---|---|---|
| | **Coeff (95% CI)** | ***P* value** | **Coeff (95% CI)** | ***P* value** |
| Non-mutation & symptomatic | -1.11 (-1.84,-0.37) | **0.003** | -1.27 (-1.42,-1.12) | **$< 2 \times 10^{-16}$** |
| GRN & presymptomatic | -0.65 (-1.74,0.44) | 0.245 | -0.07 (-0.34, 0.21) | 0.632 |
| GRN & symptomatic | -3.07 (-4.33,-1.89) | **$8.98 \times 10^{-7}$** | -1.22 (-1.47, -0.96) | **$< 2 \times 10^{-16}$** |
| C9 & presymptomatic | -1.99 (-2.80,-1.19) | **$1.68 \times 10^{-6}$** | -0.06 (-0.25, 0.12) | 0.510 |
| C9 & symptomatic | -3.49 (-4.40,-2.58) | **$1.77 \times 10^{-13}$** | -0.98 (-1.17, -0.78) | **$< 2 \times 10^{-16}$** |
| MAPT & presymptomatic | -0.68 (-1.71,0.35) | 0.199 | 0.19 (-0.07, 0.45) | 0.158 |
| MAPT & symptomatic | -2.71 (-3.77,-1.68) | **$4.72 \times 10^{-7}$** | -1.04 (-1.27, -0.80) | **$< 2 \times 10^{-16}$** |
| Education | 0.07 (-0.01,0.16) | 0.110 | 0.06 (0.04, 0.07) | **$1.15 \times 10^{-11}$** |
| Age at visit | -0.18 (-0.20,-0.16) | **$< 2 \times 10^{-16}$** | -0.02 (-0.02, -0.01) | **$< 2 \times 10^{-16}$** |
| Sex (female) | 1.75 (1.32,2.16) | **$2.14 \times 10^{-15}$** | -0.10 (-0.19, -0.02) | **0.021** |
| CDR+NACC-FTLD SB | -0.39 (-0.44,-0.33) | **$< 2 \times 10^{-16}$** | -0.11 (-0.13, -0.10) | **$< 2 \times 10^{-16}$** |

**eTable 6. Linear mixed models were fitted with education, age, sex, genetic status (non-carrier vs carrier) and the CDR+NACC-FTLD sum of boxes as fixed effects and pedigree as random effect to investigate the association between *TMEM106B* rs1990622 and the outcome measures total gray matter volume and UDS3-EF.**

| | ***Total gray matter volume (N = 958)*** | | ***UDS3-EF (N = 1581)*** | |
|---|---|---|---|---|
| | **Coeff (95% CI)** | ***P* value** | **Coeff (95% CI)** | ***P* value** |
| TMEM106B Additive | -0.08 (-0.39,0.22) | 0.602 | -0.04 (-0.10, 0.03) | 0.291 |
| TMEM106B Recessive | -0.20 (-0.78,0.37) | 0.489 | -0.05 (-0.17,0.07) | 0.457 |

**eTable 7. Linear mixed models were fitted with education, age, sex, genetic status (non-carrier vs carrier) and the CDR+NACC-FTLD sum of boxes as fixed effects and pedigree as random effect to investigate the association between *TMEM106B* rs1990622 and the outcome measures total gray matter volume and UDS3-EF in affected individuals**

| | ***Total gray matter volume N = 536*** | | ***UDS3-EF N = 1113*** | |
|---|---|---|---|---|
| | **Coeff (95% CI)** | ***P* value** | **Coeff (95% CI)** | ***P* value** |
| TMEM106B Additive | 0.01 (-0.44, 0.47) | 0.596 | 0.04 (-0.05, 0.13) | 0.338 |
| TMEM106B Recessive | -0.05 (-0.89, 0.79) | 0.910 | 0.04 (-0.11, 0.20) | 0.590 |

**eTable 8. Linear mixed model statistics for *TMEM106B* rs1990622 on individual gray matter regions of interest in *GRN* carriers with p < 0.05 in the recessive model, after controlling for years of education, sex, age at visit, and CDR+NACC-FTLD SB as fixed effects and pedigree as random effect.**

| | **Coeff (95% CI)** | ***P* value** |
|---|---|---|
| Left thalamus | 0.03 (0.01,0.06) | 0.006 |
| Right cerebellum | 0.24 (0.04,0.43) | 0.020 |
| Left cerebellum | 0.23 (0.02,0.43) | 0.035 |
| Left rostral anterior cingulate | 0.011 (0.0007,0.02) | 0.040 |
| Right thalamus | 0.03 (0.001,0.05) | 0.044 |

**eTable 9. Linear mixed model statistics for *TMEM106B* rs1990622 on individual gray matter regions of interest in presymptomatic *GRN* carriers with p < 0.05 in the recessive model, after controlling for years of education, sex and age at visit as fixed effects and pedigree as random effect**

| | **Coeff (95% CI)** | ***P* value** |
|---|---|---|
| Left thalamus | 0.03 (0.01,0.05) | 0.003 |
| Right cerebellum | 0.26 (0.08,0.44) | 0.008 |
| Right insular cortex | 0.02 (0.008,0.04) | 0.008 |
| Left temporal cortex | 0.17 (0.04,0.30) | 0.013 |
| Right frontal cortex | 0.25 (0.07,0.44) | 0.014 |
| Right temporal cortex | 0.15 (0.03,0.28) | 0.024 |
| Left frontal cortex | 0.24 (0.04,0.43) | 0.025 |
| Right thalamus | 0.03 (0.004,0.05) | 0.029 |
| Left cerebellum | 0.26 (0.03,0.49) | 0.032 |

**eTable 10. Longitudinal linear mixed statistics for the association of *TMEM106B* rs1990622 with UDS-3 in presymptomatic *C9orf72* carriers**

| | **Additive** | | **Recessive** | |
|---|---|---|---|---|
| | **Coeff (95% CI)** | ***P* value** | **Coeff (95% CI)** | ***P* value** |
| TMEM106B rs1990622 | 0.22 (0.05, 0.39) | **0.014** | 0.45 (0.13, 0.78) | **0.008** |
| Time since baseline | 0.06 (0.0001, 0.12) | 0.06 | 0.06 (-0.001, 0.12) | 0.07 |
| Education | 0.05 (-0.003, 0.11) | 0.06 | 0.05 (-0.003, 0.11) | 0.05 |
| Sex (female) | -0.003 (-0.25, 0.25) | 0.98 | -0.03 (-0.27, 0.22) | 0.82 |
| Age at baseline | -0.02 (-0.03, -0.01) | **0.0003** | -0.02 (-0.03, -0.10) | **0.0003** |
